# Observed Versus Modeled Data in GLOBOCAN 2024: Variation Across Cancer Sites and Implications for the Mortality-to-Incidence Ratio

**DOI:** 10.64898/2026.11.17.26368346

**Authors:** Srijana Dhakal, Sleshma Kalika

**Affiliations:** Department of Head and Neck Surgery, BP Koirala Memorial Cancer Hospital, Bharatpur, Chitwan, Bagmati Province, Nepal; Department of Computer Science and Information Technology, Patan Multiple Campus, Tribhuvan University, Kathmandu, Bagmati Province, Nepal

**Keywords:** GLOBOCAN, cancer incidence, cancer mortality, estimation methods, cancer registries, mortality-to-incidence ratio, Human Development Index

## Abstract

**Purpose:** The mortality-to-incidence ratio (MIR) from GLOBOCAN is often used to compare cancer outcomes across countries. But in many countries, its two components come from different sources with some estimation methods deriving one from the other. We counted the countries in which both components of MIR are observed, and tested whether correlations between MIR and the Human Development Index (HDI) hold in them.

**Methods:** Using GLOBOCAN 2024 data, we classed a country as having independent MIR when incidence came from cancer registries, mortality was observed, and neither was derived from the other. For each of 37 cancer sites, we calculated the fraction of global new cases occurring in these countries. We compared MIR-HDI Spearman correlations (ρ) in these countries with the full sample, testing for attenuation beyond range restriction against an HDI-matched null distribution.

**Results:** Only 41 of 186 countries (22%), all with high or very high HDI, had independent MIR. Of the 144 countries with derived MIR, 131 relied on external models. The independent-MIR fraction ranged from 0.12 (Kaposi sarcoma) to 0.77 (thyroid). For 33 of 37 sites, the correlation in independent-MIR countries was within what the narrower HDI range alone would produce. After Benjamini-Hochberg correction, thyroid was the only site whose correlation weakened more than the narrower HDI range would explain (ρ −0.82 to −0.004, adjusted p = 0.011).

**Conclusion:** No low- or medium-HDI country has independent MIR, and in 131 countries at least one MIR component comes from an external model. Comparing MIR across these countries mostly compares model inputs. In independent-MIR countries, most MIR-HDI associations are consistent with range restriction but for thyroid, the association disappears.

## Introduction

The 2024 GLOBOCAN edition provides estimates of global cancer incidence and mortality covering 34 cancers (37 individual sites in the database) and 186 countries [1]. The estimates are constructed using various data sources and statistical methods. For incidence, some countries contribute data from national or subnational cancer registries that directly count cases, while others rely on models that estimate incidence from mortality data, survival statistics, or by using references from neighboring countries [2,3]. The GLOBOCAN 2024 edition uses the same estimation method hierarchy as described for the 2022 edition [1,3], with incidence classified across eight method levels (1, 2a, 2b, 3a, 3b, 3c, 4, 5) and mortality across seven method levels (1, 2a, 2b, 3a, 3b, 3c, 4). For mortality, some countries record cancer deaths through vital registration systems or cancer registries, while others have mortality estimated from incidence using survival-based models [3]. In the 2022 edition, complete national incidence rates were available for only 55 countries (30%) [4]. Such discrepancy aligns closely with economic development, as only 22% of low-income countries maintain a national cancer registry compared to 75% of high-income countries [5].

While such literature on country-level variation in data sources is available, variation of estimation methods across cancer sites is understudied. Because GLOBOCAN applies a single estimation method to all cancer types within a given country, the proportion of observed versus modeled estimates for any specific cancer depends on its geographic distribution. Also, since incidence and mortality methods are assigned independently, the two components of the mortality-to-incidence ratio (MIR^1^) can have different data origins within the same country. Two prior studies partially addressed this question for individual cancers. Comparison against Norwegian registry data demonstrated that estimation accuracy varies by anatomical site [6], and site-specific analyses of gallbladder cancer revealed discrepancies between national modeled estimates and subnational registry data [7]. However, neither study systematically quantified estimation method composition across all sites, nor examined the mortality component. MIR is widely used as a proxy for cancer outcomes and health system performance [8–13], and patterns of MIR in relation to the Human Development Index (HDI) have been described for breast [14], thyroid [15], cervical [10], colorectal [11], and lip and oral cavity [12] cancers, though its validity as a survival proxy has been tested with mixed results [16,17].

Some GLOBOCAN methods derive one MIR component from the other. Mortality method 3c (85 countries) computes mortality from incidence as MIR = 1 - (5-year survival), with survival taken from one of three alternative models based on SURVCAN-3 data and stratified by world area, continent, or a two-tier HDI grouping [18]. Incidence methods 3a to 3c (51 countries) derive incidence from mortality, and method 3b (17 countries for incidence or mortality) scales the ratios by HDI [2,18]. In these countries MIR largely reproduces the ratio used to derive the missing component, and correlating it with HDI could potentially introduce circularity. Some studies relating to MIR restrict their samples to those with better data availability or quality [10,12,13], but, to our knowledge, none has classified countries by whether their MIR is independently observed.

Our main question was whether MIR-HDI correlations change when the analysis is restricted to countries where MIR is independently observed, beyond what the narrower HDI range of those countries would produce. To answer it, we first classified every country by whether its MIR is independent, and described, for each of the 37 sites, how much of the global case count comes from countries with observed incidence, mortality, and MIR.

## Methods

### Data Sources

This was an ecological study of publicly available secondary data from GLOBOCAN 2024. Data were obtained from the Global Cancer Observatory (GCO) website in September 2026 [19]. We extracted incidence and mortality estimates for 186 countries and 37 cancer sites, each country’s estimation method codes, and HDI values from the Human Development Report 2023-2024 [20]. Method codes were retrieved from the GCO application programming interface and matched with the country table in Annex A of the GLOBOCAN 2024 documentation for all 186 countries [18]. For nine territories lacking HDI values, the HDI of the administering country was used. The Democratic People’s Republic of Korea was excluded from HDI-dependent analyses due to missing HDI data. Non-melanoma skin cancer and aggregate categories were excluded. Two of the 37 sites, "other specified cancers" and "unspecified sites", are residual categories.

### Estimation Method Classification

Incidence methods 1, 2a, and 2b were classed as registry-based (90 countries) and methods 3a to 5 as modeled (96 countries). Mortality methods 1, 2a, and 2b were classed as observed (89 countries) and methods 3a to 4 as modeled (97 countries) (Table 1). Observed mortality comes from vital registration in all countries except China which is the only country with mortality method 2b (cancer registry mortality) [18,19]. A country was deemed to have independent MIR when both components were observed and neither was derived from the other (41 countries), and derived MIR when at least one component was derived from the other (144 countries). In China both components are weighted averages from the same system of subnational cancer registries [18], so we report case-weighted results with and without China. Within the derived group, 13 countries use their own registry-derived ratios (for example, India for mortality) [18], and 131 rely on external models (methods 3b, 3c, or regional or neighboring-country estimates). For the own-data group, MIR reflects locally observed data from a different period or subpopulation rather than an external model. South Africa (incidence method 4, mortality method 1) was classified separately.

**Table 1.** GLOBOCAN 2024 Estimation Methods and Country Counts.

| Component | Method | Description | No. of Countries | Classification in this study |
|---|---|---|---|---|
| Incidence | 1 | Trends in national (or >70% coverage) registry rates with >6 years of recent data, projected to 2024 | 39 | Registry-based |
|  | 2a | Recent national registry rates applied to the 2024 population | 18 | Registry-based |
|  | 2b | Recent rates from one or more subnational registries applied to the 2024 population | 33 | Registry-based |
|  | 3a | National mortality with M:I ratios from the country's own cancer registries | 11 | Modeled (own-data ratio) |
|  | 3b | National mortality with M:I ratios from neighboring countries' registries, scaled by HDI | 11 | Modeled (external; HDI-scaled) |
|  | 3c | National mortality with M:I ratios based on SURVCAN-3 survival data | 29 | Modeled (external) |
|  | 4 | Local rates scaled to all-cancer rates averaged from national estimates or neighboring countries | 12 | Modeled (borrowed) |
|  | 5 | Average rates from neighboring countries | 33 | Modeled (borrowed) |
| Mortality | 1 | National vital registration with sufficient recorded deaths, projected to 2024 | 84 | Observed |
|  | 2a | Recent vital registration rates used as proxy for 2024 | 4 | Observed |
|  | 2b | Recent cancer registry mortality used as proxy for 2024 (China) | 1 | Observed |
|  | 3a | National incidence with I:M ratios from the country's own cancer registries | 2 | Modeled (own-data ratio) |
|  | 3b | National incidence with ratios from neighboring countries, scaled by HDI | 6 | Modeled (external; HDI-scaled) |
|  | 3c | National incidence with M:I = 1 − 5-year survival from one of three SURVCAN-3-based models (stratified by world area, continent, or two-tier HDI); NORDCAN correction factor for 14 sites | 85 | Modeled (external) |
|  | 4 | Rates scaled to all-cancer mortality averaged from national estimates or neighboring countries | 4 | Modeled (borrowed) |

*NOTE. Descriptions are summarized from the GLOBOCAN 2024 annexes. The NORDCAN correction applies to oropharynx, esophagus, colon, liver, gallbladder, pancreas, breast, ovary, vulva, kidney, brain/CNS, thyroid, multiple myeloma, and leukemia.*

### Estimation Method Composition

For each of the 37 cancer sites, we calculated the proportion of total global estimated new cases (both sexes combined) derived from countries classified under each estimation method category. We define three summary metrics:

(i) the incidence observed fraction (proportion from registry-based countries);

(ii) the mortality observed fraction (proportion from vital registration countries); and

(iii) the fully observed MIR fraction (proportion from countries where both are empirically observed).

Cancer sites were ranked by ascending incidence observed fraction. We evaluated the Spearman rank correlation between the incidence observed fraction and the proportion of cases in low-HDI and medium-HDI countries (HDI < 0.70 is the UNDP threshold between medium and high).

### MIR-HDI Analysis

Country MIR was computed for each site as the ratio of the age-standardized mortality rate to the age-standardized incidence rate, excluding records with fewer than 100 estimated cases or an incidence rate of zero. For each site, we calculated the Spearman rank correlation (ρ) between MIR and HDI across all eligible countries (full sample), then repeated it under four restriction schemes:

Scheme A. Incidence-observed only: restricting to countries with registry-based incidence (standardizing the MIR denominator).

Scheme B. Mortality-observed only: restricting to countries with vital registration-based mortality (standardizing the MIR numerator).

Scheme C. Fully observed: restricting to countries with independent MIR.

Scheme D. Method-3b exclusion: excluding only the countries using incidence method 3b to isolate potential HDI-based circularity.

The change in correlation was summarized as Δρ = ρ_restricted – ρ_full, with 95% Confidence Intervals (CIs) from 2,000 paired bootstrap resamples. Because most correlations were negative, a positive Δρ means attenuation. Across sites, we report the median absolute Δρ, which ignores the direction of change.

Since the restricted samples are concentrated at high HDI, some attenuation is expected simply because of the narrower range of HDI i.e., range restriction alone. To separate this from an effect of estimation method, we drew 10,000 random subsets of distinct countries from the full sample for Schemes A to C. Each subset had the same number of countries in each HDI quartile as the restricted sample. The correlations in these subsets show what the narrower HDI range alone would produce, regardless of estimation method. We call this the HDI-matched null distribution, and its central 95% the null interval. The one-sided p-value was the proportion of subsets with a correlation at least as close to zero as the observed value, computed as (k+1)/(N+1), and p-values were adjusted across the 37 sites with the Benjamini-Hochberg (BH) correction.

### Influence and Sensitivity Analyses

For sites with BH-significant attenuation, we removed each Scheme C country in turn and recomputed the correlation (leave-one-out analysis). We also repeated the subsampling with HDI deciles instead of quartiles, which matches the HDI distribution more closely. Further sensitivity analyses excluded China and India and counted incidence method 3a as registry-based.

All analyses were done using Python 3.13 (*pandas, NumPy, SciPy*) and Microsoft Excel. The visualizations were made with R. The study used publicly available, aggregated, country-level data, therefore did not require ethics committee approval.

## Results

### Country Classification

Of 186 countries, 90 (48.4%) had registry-based incidence estimates (method 1: n = 39, method 2a: n = 18, method 2b: n = 33) and 96 (51.6%) had modeled incidence estimates. For mortality, 89 countries (47.8%) had vital registration-based estimates (method 1: n = 84; method 2a: n = 4; method 2b: n = 1) while 97 (52.2%) had modeled mortality estimates of which 85 used survival-based modeling (method 3c). In many countries only one of the two was observed: 49 countries had registry-based incidence but modeled mortality, and 48 had the opposite (Table 2). Only 41 countries (22.0%) had independent MIR. None of them had low or medium HDI (35 very high, 6 high). Of the 144 with derived MIR, 13 used their own registry-derived ratios and 131 relied on external models (Supplementary Table S1).

**Table 2.** Status of incidence and mortality estimation methods in GLOBOCAN 2024.

|  | **Mortality observed (methods 1, 2a, 2b)** | **Mortality modeled (methods 3a–4)** | Total |
|---|---|---|---|
| **Incidence observed (methods 1, 2a, 2b)** | 41 | 49 | 90 |
| **Incidence modeled (methods 3a–5)** | 48 | 48 | 96 |
| Total | 89 | 97 | 186 |

### Estimation Method Composition Across Cancer Sites

The incidence observed fraction ranged from 0.55 (Kaposi sarcoma) to 0.87 (thyroid) with a median of 0.74 (Figure 1, Table 3). The five cancer sites with the lowest fractions were Kaposi sarcoma (0.55), testis (0.62), Hodgkin lymphoma (0.65), cervix uteri (0.67), and anus (0.68). Borrowed incidence (methods 4 and 5) made up the largest share for Kaposi sarcoma (26.7%), nasopharynx (22.6%), and cervix uteri (18.7%).

**Figure 1.**
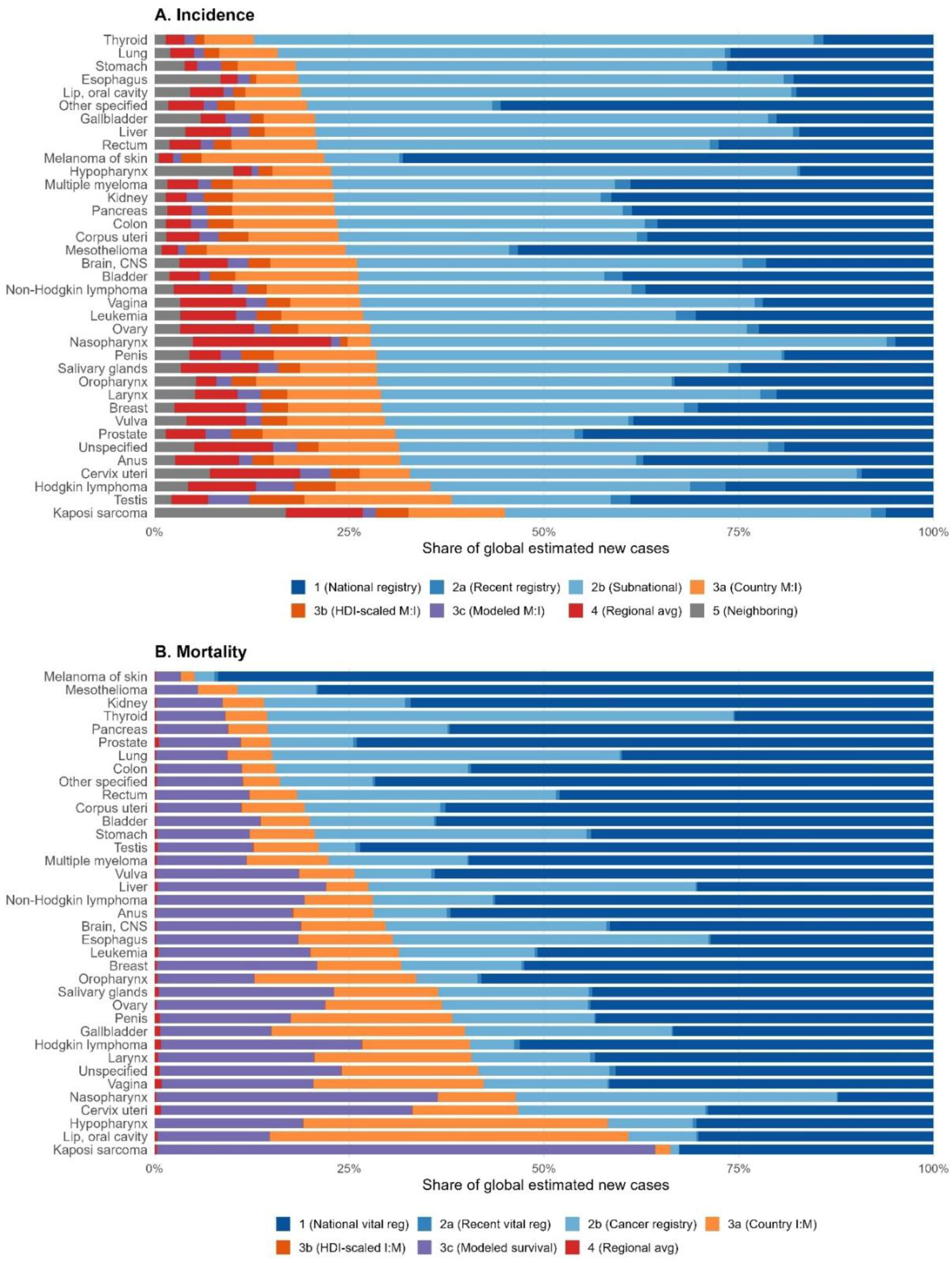
Estimation method composition for 37 cancer sites in GLOBOCAN 2024, weighted by estimated new cases. (A) Incidence estimation methods. (B) Mortality estimation methods. Sites are ordered from highest (top) to lowest (bottom) observed fraction. Blue shades: observed methods (registry-based for incidence, vital registration or cancer registry for mortality; methods 1, 2a, 2b). Orange shades: methods 3a and 3b. Purple: method 3c. Red: method 4 (regional averages). Gray: method 5 (neighboring countries, incidence only).

**Table 3.** Estimation method composition by cancer site in GLOBOCAN 2024.

| **Cancer site** | **Total estimated cases** | **Incidence observed fraction** | **Mortality observed fraction** | **Independent-MIR fraction** |
|---|---|---|---|---|
| Kaposi sarcoma | 34,205 | 0.550 | 0.338 | 0.120 |
| Testis | 70,658 | 0.618 | 0.789 | 0.477 |
| Hodgkin lymphoma | 84,417 | 0.645 | 0.595 | 0.371 |
| Cervix uteri | 603,890 | 0.672 | 0.533 | 0.384 |
| Anus | 56,389 | 0.685 | 0.719 | 0.502 |
| Unspecified sites | 470,328 | 0.686 | 0.584 | 0.420 |
| Prostate | 1,544,523 | 0.691 | 0.851 | 0.603 |
| Vulva | 48,874 | 0.704 | 0.743 | 0.541 |
| Breast | 2,432,225 | 0.708 | 0.683 | 0.506 |
| Larynx | 193,774 | 0.709 | 0.593 | 0.410 |
| Oropharynx | 107,835 | 0.714 | 0.664 | 0.458 |
| Salivary glands | 55,212 | 0.715 | 0.636 | 0.485 |
| Penis | 34,755 | 0.715 | 0.618 | 0.411 |
| Nasopharynx | 119,725 | 0.722 | 0.536 | 0.486 |
| Ovary | 330,532 | 0.723 | 0.631 | 0.480 |
| Leukemia | 494,787 | 0.732 | 0.686 | 0.524 |
| Vagina | 18,139 | 0.735 | 0.577 | 0.423 |
| Non-Hodgkin lymphoma | 562,345 | 0.737 | 0.720 | 0.555 |
| Bladder | 634,723 | 0.738 | 0.801 | 0.596 |
| Brain, central nervous system | 323,901 | 0.740 | 0.703 | 0.537 |
| Mesothelioma | 27,333 | 0.755 | 0.893 | 0.670 |
| Corpus uteri | 434,260 | 0.763 | 0.807 | 0.627 |
| Colon | 1,205,148 | 0.765 | 0.844 | 0.656 |
| Pancreas | 530,979 | 0.769 | 0.855 | 0.670 |
| Kidney | 442,274 | 0.770 | 0.860 | 0.670 |
| Multiple myeloma | 195,993 | 0.771 | 0.776 | 0.600 |
| Hypopharynx | 91,455 | 0.773 | 0.418 | 0.315 |
| Melanoma of skin | 337,714 | 0.782 | 0.949 | 0.749 |
| Rectum | 778,207 | 0.791 | 0.817 | 0.665 |
| Liver and intrahepatic bile ducts | 842,670 | 0.794 | 0.725 | 0.619 |
| Gallbladder | 126,349 | 0.794 | 0.602 | 0.492 |
| Other specified cancers | 1,869,342 | 0.804 | 0.839 | 0.703 |
| Lip, oral cavity | 452,073 | 0.812 | 0.392 | 0.293 |
| Esophagus | 494,147 | 0.815 | 0.694 | 0.613 |
| Stomach | 979,869 | 0.819 | 0.794 | 0.669 |
| Trachea, bronchus and lung | 2,635,551 | 0.842 | 0.849 | 0.741 |
| Thyroid | 958,835 | 0.873 | 0.855 | 0.767 |

*NOTE. Sites ranked by ascending incidence observed fraction. Each fraction is the share of global estimated new cases (both sexes) that occur in countries with registry-based incidence (methods 1, 2a, 2b), observed mortality (methods 1, 2a, 2b), or independent MIR.*

The distribution of mortality estimation methods exhibited a bimodal pattern (Figure 1), dominated by vital registration (method 1: n = 84 countries) and survival-derived modeling (method 3c: n = 85 countries). The mortality observed fraction ranged from 0.34 for Kaposi sarcoma to 0.95 for melanoma (median: 0.72). This does not mirror the incidence fraction. For instance, lip and oral cavity had the fifth-highest incidence observed fraction (0.81) but has among the lowest mortality observed fractions (0.39), reflecting countries (such as India) that maintain cancer registries but lack complete vital registration.

The independent-MIR fraction ranged from 0.12 (Kaposi sarcoma) to 0.77 (thyroid) with a median of 0.54 (Table 3). Overall, 12.6 of 20.6 million estimated cases (61%) occurred in independent-MIR countries, falling to 48% when China (25% of global cases) was excluded. Across sites, a larger share of cases in low-HDI and medium-HDI countries was associated with a lower incidence observed fraction (ρ = −0.41; ρ = −0.74 when method 3a was counted as observed).

Restricting to observed mortality (Scheme B, 89 countries) resulted in larger changes, mostly toward zero (median absolute Δρ = 0.076; 29 of 37 sites attenuated), significant only for gallbladder (BH p = 0.033). Excluding method-3b countries (Scheme D) had negligible effect (median absolute Δρ = 0.008) (Supplementary Table S2).

Under the independent-MIR restriction (Scheme C, 41 countries), the median absolute Δρ was 0.132. These countries had higher HDI with narrower range (median 0.91, IQR 0.82-0.94) than the full sample (median 0.74, IQR 0.60-0.88).

For 33 of 37 sites, the Scheme C correlation was within the null interval, so its change can be explained by the narrower HDI range alone (Figure 2, Supplementary Table S3). The null intervals were wide (median width 0.29 units of ρ, with a median of 33 countries per site), so only changes much larger than expected from the narrower HDI range could be detected. Seven sites had nominal one-sided p < 0.05, and four of these (thyroid, ovary, kidney, and brain/CNS) fell outside the null interval. After BH correction, only thyroid showed attenuation beyond range restriction (BH p = 0.011). The next-strongest attenuation, for ovary (ρ −0.72 in the full sample and ρ = −0.02 under Scheme C), was not significant after BH correction (BH p = 0.056).

**Figure 2.**
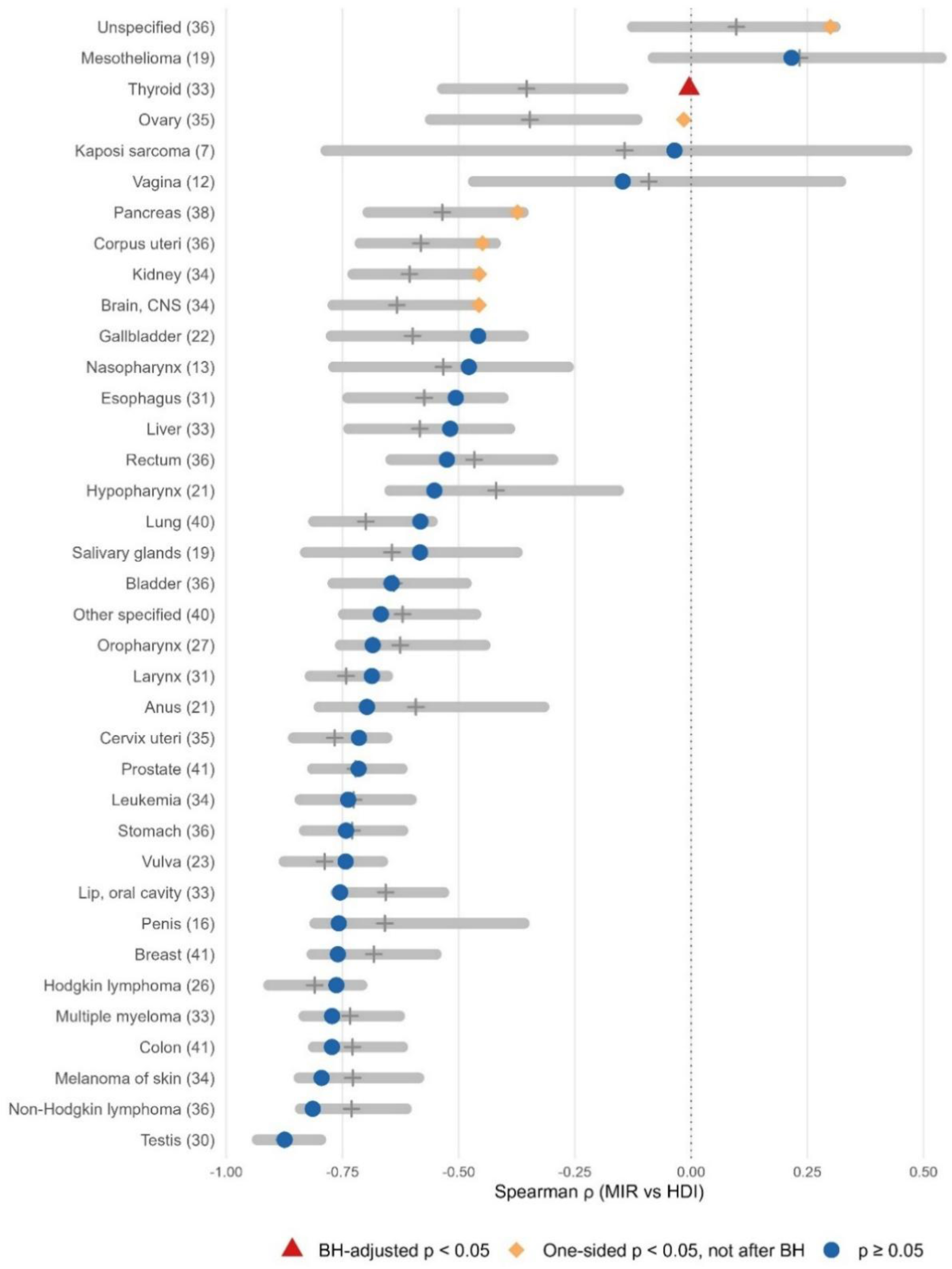
Spearman correlation between MIR and HDI in independent-MIR countries for each cancer site (number of countries in parentheses), against the HDI-matched null distribution. Gray bars: null interval (central 95% of correlations in random HDI-matched subsets). Crosses: null median. Red triangle: attenuation beyond range restriction after Benjamini-Hochberg correction. Orange diamonds: one-sided p < 0.05 but not after correction. Blue circles: one-sided p ≥ 0.05. Sites are ordered by observed correlation.

For thyroid, the correlation fell from −0.82 in the full sample to −0.004 in independent-MIR countries, outside the null interval (-0.53 to −0.15, median −0.35) (Figure 3A).

**Figure 3.**
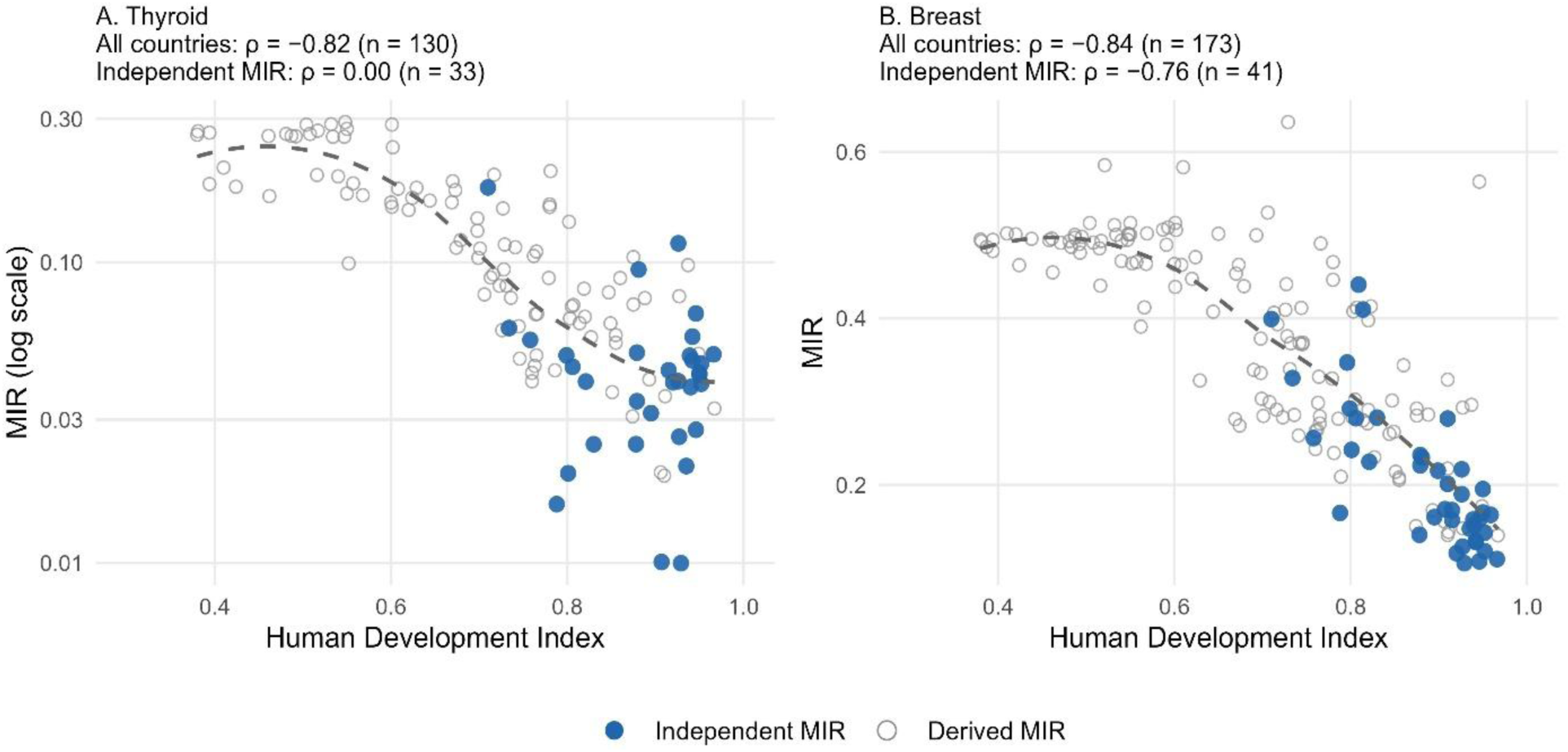
MIR versus HDI for thyroid cancer (A, log scale) and breast cancer (B). Each point is a country. Blue filled circles: countries with independent MIR. Gray open circles: countries with derived MIR. Gray dashed line: Local regression fit for all countries. For thyroid MIR is not correlated with HDI in independent-MIR countries (ρ = −0.004), while the full sample shows a strong negative correlation (ρ = −0.82). For breast, the correlation sustains in independent-MIR countries (ρ = −0.76).

Neither Scheme A (p = 0.10) nor Scheme B (p = 0.59) alone produced significant attenuation. Removing any one country did not restore the correlation (leave-one-out ρ, −0.06 to 0.09), and matching on HDI deciles instead of quartiles had similar result (p = 0.0003). Breast cancer kept its correlation in independent-MIR countries (ρ = −0.76, compared with ρ = −0.84 in the full sample; Figure 3B).

Excluding China and India lowered the incidence observed fraction by a median of 0.11 across sites (nasopharynx, 0.72 to 0.44) but changed full-sample MIR-HDI correlations by a median of only 0.006, and counting method 3a as registry-based gave results close to Scheme A (median absolute Δρ = 0.028 vs 0.024).

## Discussion

In 144 of 186 countries (77%), MIR is not an independent measurement, because one of its components is derived from the other, and in 131 of them, this involves an external model. None of the 41 countries with independent MIR has low or medium HDI, and the share of each cancer’s estimated new cases occurring in independent-MIR countries ranges from 0.12 for Kaposi sarcoma to 0.77 for thyroid. Within the HDI range that can be checked, most MIR-HDI correlations are consistent with range restriction except for thyroid.

Classifying estimation methods for both components shows that the incidence and mortality parts of MIR often come from different kinds of data. 49 countries have registry-based incidence with modeled mortality, and 48 have the opposite. For the 85 countries using method 3c, MIR is a function of the survival assumed for that country rather than an observed ratio. Although 77% of countries have derived MIR, they account for only 39% of global cases, because a few countries with very many cases, such as China, have independent MIR. Restricting samples by data availability or quality [10,12] removes many derived-MIR countries, but because observed data are concentrated at high HDI, such samples face the same range restriction as Scheme C. Observed data tend to be scarcer for cancers whose cases are concentrated in lower-HDI countries (ρ = −0.41 across sites). For cervix uteri, about a third of the roughly 604,000 estimated cases come from countries with modeled incidence (observed fraction 0.67), and only 38% come from countries with independent MIR. Of the 151 countries in our cervical cancer full sample, 116 had derived MIR.

Within the high-HDI range where independent data exist, deriving one MIR component from the other did not noticeably change MIR-HDI correlations for most sites. Excluding the method-3b countries, where HDI enters the model directly, also had negligible effect. MIR-HDI associations at low and medium HDI cannot be checked against independent data, because no low- or medium-HDI country has independent MIR. Thyroid is an exception. Across all countries, thyroid MIR falls steeply as HDI rises (ρ - 0.82), but this correlation disappeared in independent-MIR countries. It disappeared only when both components were restricted, and the attenuation did not depend on any single country. In the 85 method-3c countries, thyroid mortality is computed from incidence using a mortality-to-incidence ratio set to one minus modeled 5-year survival, and then adjusted with a NORDCAN-based correction factor, which GLOBOCAN applies for thyroid and 13 other sites [18,21]. The survival models are stratified by world area, continent, or two-tier HDI group, so modeled MIR in these countries varies with these groupings by design. Thyroid incidence also depends on how intensively the thyroid is examined. Increased use of imaging finds small, subclinical tumors that would not have caused symptoms or death, and the proportion of cases attributed to overdiagnosis varies widely between countries [22]. Detecting such tumors raises incidence without necessarily adding deaths and so lowers MIR [15,22]. In countries with independent MIR, observed thyroid MIR may therefore largely reflect how intensively tumors are detected, which need not follow HDI. All four sites whose correlation fell outside the null interval (thyroid, ovary, kidney, and brain/CNS) are among the 14 NORDCAN-corrected sites. Breast is also one of the 14 but kept its correlation, so the correction alone does not explain the pattern. These observations are consistent with, but do not establish, an effect of the survival model.

## Limitations

GLOBOCAN assigns methods at the national level, so differences in registry coverage between cancers within a country are not captured, and the published documentation does not state which survival model applies to each method-3c country. Method 2b covers a wide range of subnational coverage, and China, classified as method 2b for both components and accounting for 25% of global cases, strongly influences case-weighted fractions. Our classification also counts the 13 method-3a countries as modeled, although they use their own registry data rather than external models.

Counting them as observed would raise the incidence observed fraction by a median of 0.12 across sites (up to 0.19 for testis), so the incidence observed fractions reported here are with a conservative stance. Also, the independent-MIR sample is small, six sites had fewer than 20 eligible countries, and therefore, the test could detect only large effects of estimation method.

## Recommendations

MIR-HDI analyses should report independent-MIR and derived-MIR countries separately, for example with different plot symbols as in Figure 3, and should state that associations at low and medium HDI cannot yet be checked against independent data. Researchers comparing thyroid MIR across development levels should exercise particular caution. Stating in GLOBOCAN documentation which survival model is applied to each method-3c country would show users where modeled MIR is tied to HDI. The 49 countries that already have registry-based incidence but modeled mortality are where improved vital registration, which has been slow to develop in many low- and medium-HDI settings [23], would most directly increase the number of independent-MIR countries.

In conclusion, MIR is independently measured in only 41 of the 186 countries in GLOBOCAN 2024, and in none with low or medium HDI. In the other 144 countries (77%), comparing MIR between countries largely compares the ratios used in estimation. In the 41 independent-MIR countries, most MIR-HDI associations are consistent with range restriction, but the thyroid association disappears. The share of each cancer’s cases occurring in independent-MIR countries also varies, from 0.12 for Kaposi sarcoma to 0.77 for thyroid.

## Author Contributions

Conception and design: Srijana Dhakal. Collection and assembly of data: Sleshma Kalika. Data analysis and interpretation: Both authors. Manuscript writing: Both authors. Final approval of manuscript: Both authors.

## Data Availability

All data used in this study are publicly available from the Global Cancer Observatory (https://gco.iarc.who.int/today/) and the UNDP Human Development Reports (https://hdr.undp.org/). The country MIR classification for all 186 countries is provided in the Data Supplement (Table S1) and as a CSV file.

## Footnotes

1 MIR = Age-Standardized Mortality Rate (ASMR) / Age-Standardized Incidence Rate (ASIR)

## References

1. Sung H, Filho AM, Laversanne M, Ferlay J, Siegel RL, Soerjomataram I, et al. Global cancer statistics 2024: GLOBOCAN estimates of incidence and mortality worldwide for 34 cancers in 186 countries. CA A Cancer J Clinicians. 2026 Jul;76(4):e70090. doi:10.3322/caac.70090

2. Ferlay J, Colombet M, Soerjomataram I, Mathers C, Parkin DM, Piñeros M, et al. Estimating the global cancer incidence and mortality in 2018: GLOBOCAN sources and methods. Intl Journal of Cancer. 2019 Apr 15;144(8):1941–53. doi:10.1002/ijc.31937

3. Filho AM, Laversanne M, Ferlay J, Colombet M, Piñeros M, Znaor A, et al. The GLOBOCAN 2022 cancer estimates: Data sources, methods, and a snapshot of the cancer burden worldwide. Intl Journal of Cancer. 2025 Apr;156(7):1336–46. doi:10.1002/ijc.35278

4. Hemminki K, Kaaks R. The new GLOBOCAN methods paper demonstrates global inequalities in data sources. Intl Journal of Cancer. 2025 Apr;156(7):1313–1313. doi:10.1002/ijc.35300

5. Siddiqui AH, Zafar SN. Global Availability of Cancer Registry Data. JGO. 2018 Jun;4:1–3. doi:10.1200/JGO.18.00116

6. Antoni S, Soerjomataram I, Møller B, Bray F, Ferlay J. An assessment of GLOBOCAN methods for deriving national estimates of cancer incidence. Bull World Health Organ. 2016 Mar 1;94(3):174–84. doi:10.2471/BLT.15.164384

7. Piñeros M, Vignat J, Colombet M, Laversanne M, Ferreccio C, Heise K, et al. Global variations in gallbladder cancer incidence: What do recorded data and national estimates tell us? Intl Journal of Cancer. 2025 Apr;156(7):1358–68. doi:10.1002/ijc.35232

8. Choi E, Lee S, Nhung BC, Suh M, Park B, Jun JK, et al. Cancer mortality-to-incidence ratio as an indicator of cancer management outcomes in Organization for Economic Cooperation and Development countries. Epidemiol Health. 2017 Feb 5;39:e2017006. doi:10.4178/epih.e2017006

9. Sunkara V, Hébert JR. The colorectal cancer mortality-to-incidence ratio as an indicator of global cancer screening and care. Cancer. 2015 May 15;121(10):1563–9. doi:10.1002/cncr.29228

10. Shen TT, Long CY, Wu MP. Favorable cervical cancer mortality-to-incidence ratios of countries with good human development index rankings and high health expenditures. BMC Women’s Health. 2023 May 25;23(1):284. doi:10.1186/s12905-023-02423-y

11. Wang CC, Sung WW, Yan PY, Ko PY, Tsai MC. Favorable colorectal cancer mortality-to-incidence ratios in countries with high expenditures on health and development index: A study based on GLOBOCAN database. Medicine. 2021 Oct 15;100(41):e27414. doi:10.1097/MD.0000000000027414

12. Sung WW, Hsu YC, Dong C, Chen YC, Chao YC, Chen CJ. Favorable Lip and Oral Cancer Mortality-to-Incidence Ratios in Countries with High Human Development Index and Expenditures on Health. Int J Environ Res Public Health. 2021 Jun 3;18(11):6012. doi:10.3390/ijerph18116012 PubMed PMID: 34205053; PubMed Central PMCID: PMC8199886.

13. Diehl TM, Ahmed KS, Pourdashti S, Stalter L, Hellner J, Harrison E, et al. Disparities in cancer mortality worldwide: a novel metric for measuring global disparities and prioritizing cancer control efforts. JCO Global Oncology. 2025;11:e2400336. doi:10.1200/GO-24-00336

14. Gu H, Wang R, Beeraka NM, Lu P, Zhao X, Li L, et al. Global burden and trends of breast cancer: GLOBOCAN 2022 estimates of incidence and mortality in 185 countries. Chin Med J (Engl). 2026 Feb 5;139(3):404–14. doi:10.1097/CM9.0000000000003921 PubMed PMID: 41473983; PubMed Central PMCID: PMC12875702.

15. Hsu YC, Cheng SY, Chien MN, Cheng S. Impact of social and economic factors on global thyroid cancer incidence and mortality. European Archives of Oto-Rhino-Laryngology. 2023;280:4185–93. doi:10.1007/s00405-023-07992-0

16. Asadzadeh Vostakolaei F, Karim-Kos HE, Janssen-Heijnen MLG, Visser O, Verbeek ALM, Kiemeney LALM. The validity of the mortality to incidence ratio as a proxy for site-specific cancer survival. European Journal of Public Health. 2011 Oct;21(5):573–7. doi:10.1093/eurpub/ckq120

17. Ellis L, Belot A, Rachet B, Coleman MP. The Mortality-to-Incidence Ratio Is Not a Valid Proxy for Cancer Survival. JGO. 2019 May;5:1–9. doi:10.1200/JGO.19.00038

18. International Agency for Research on Cancer. GLOBOCAN 2024 annexes A–C, and Annex A: Cancer incidence and mortality data sources and methods by country [Technical Report] [Internet]. Lyon, France: International Agency for Research on Cancer; 2026. Available from: https://gco.iarc.who.int/media/globocan/material/GLOBOCAN2024_annexes.pdf

19. Ferlay J, Ervik M, Lam F, Laversanne M, Colombet M, Mery L, et al. Global cancer observatory: Cancer today (GLOBOCAN 2024, data version 08.07.2026) [Internet]. Lyon, France: International Agency for Research on Cancer; 2026. Available from: https://gco.iarc.who.int/today/

20. Human Development Index. Human Development Reports [Internet]. United Nations Development Programme; [cited 2026 Sep 1]. Available from: https://hdr.undp.org/data-center/human-development-index

21. Larønningen S, Arvidsson G, Bray F, Dahl-Olsen ED, Engholm G, Ervik M, et al. NORDCAN: Cancer Incidence, Mortality, Prevalence and Survival in the Nordic Countries, Version 9.5 [Internet]. Oslo, Norway: Association of the Nordic Cancer Registries & Cancer Registry of Norway; 2025. Available from: https://nordcan.iarc.fr/

22. Li M, Dal Maso L, Pizzato M, Vaccarella S. Evolving epidemiological patterns of thyroid cancer and estimates of overdiagnosis in 2013–17 in 63 countries worldwide: a population-based study. The Lancet Diabetes & Endocrinology. 2024;12(11):824–36. 10.1016/s2213-8587(24)00223-7

23. Bray F, Znaor A, Cueva P, Korir A, Swaminathan R, Ullrich A, et al. Planning and Developing Population-Based Cancer Registration in Low- and Middle-Income Settings. Lyon, France: International Agency for Research on Cancer; 2014. (IARC Technical Publications; no. 43). PubMed PMID: 33502836.

