## Supplementary material for "Observed Versus Modeled Data in GLOBOCAN 2024: Variation Across Cancer Sites and Implications for the Mortality-to-Incidence Ratio": Data Supplement

***Supplementary Table S1.*** *MIR Classification of All 186 Countries*

| Country | ISO3 | Incidence method | Mortality method | HDI | MIR class | Derived component | Ratio source | Method 3b |
|---|---|---|---|---|---|---|---|---|
| Afghanistan | AFG | 5 | 3c | 0.462 | derived | mortality | external model | |
| Albania | ALB | 2a | 3b | 0.789 | derived | mortality | external model | Yes |
| Algeria | DZA | 2b | 3c | 0.745 | derived | mortality | external model | |
| Angola | AGO | 2b | 3c | 0.591 | derived | mortality | external model | |
| Argentina | ARG | 3a | 1 | 0.849 | derived | incidence | own-data ratio | |
| Armenia | ARM | 3c | 1 | 0.786 | derived | incidence | external model | |
| Australia | AUS | 1 | 1 | 0.946 | independent | – | – | |
| Austria | AUT | 1 | 1 | 0.926 | independent | – | – | |
| Azerbaijan | AZE | 3c | 2a | 0.760 | derived | incidence | external model | |
| Bahamas | BHS | 3c | 1 | 0.820 | derived | incidence | external model | |
| Bahrain | BHR | 1 | 3c | 0.888 | derived | mortality | external model | |
| Bangladesh | BGD | 5 | 3c | 0.670 | derived | mortality | external model | |
| Barbados | BRB | 2a | 1 | 0.809 | independent | – | – | |
| Belarus | BLR | 1 | 2a | 0.801 | independent | – | – | |
| Belgium | BEL | 1 | 1 | 0.942 | independent | – | – | |
| Belize | BLZ | 3c | 1 | 0.700 | derived | incidence | external model | |
| Benin | BEN | 2b | 3c | 0.504 | derived | mortality | external model | |
| Bhutan | BTN | 2a | 3c | 0.681 | derived | mortality | external model | |
| Bolivia (Plurinational State of) | BOL | 3c | 4 | 0.698 | derived | incidence | external model | |
| Bosnia Herzegovina | BIH | 3a | 1 | 0.779 | derived | incidence | own-data ratio | |
| Botswana | BWA | 4 | 3c | 0.708 | derived | mortality | external model | |
| Brazil | BRA | 3a | 1 | 0.760 | derived | incidence | own-data ratio | |
| Brunei Darussalam | BRN | 1 | 3c | 0.823 | derived | mortality | external model | |
| Bulgaria | BGR | 1 | 1 | 0.799 | independent | – | – | |
| Burkina Faso | BFA | 4 | 3c | 0.438 | derived | mortality | external model | |
| Burundi | BDI | 5 | 3c | 0.420 | derived | mortality | external model | |
| Cambodia | KHM | 5 | 3c | 0.600 | derived | mortality | external model | |
| Cameroon | CMR | 4 | 3c | 0.587 | derived | mortality | external model | |
| Canada | CAN | 1 | 1 | 0.935 | independent | – | – | |

| Country | ISO3 | Incidence method | Mortality method | HDI | MIR class | Derived component | Ratio source | Method 3b |
|---|---|---|---|---|---|---|---|---|
| Cape Verde | CPV | 2a | 3c | 0.661 | derived | mortality | external model | |
| Central African Republic | CAF | 5 | 3c | 0.387 | derived | mortality | external model | |
| Chad | TCD | 2b | 3c | 0.394 | derived | mortality | external model | |
| Chile | CHL | 3c | 1 | 0.860 | derived | incidence | external model | |
| China | CHN | 2b | 2b | 0.788 | independent | – | – | |
| Colombia | COL | 2b | 1 | 0.758 | independent | – | – | |
| Comoros | COM | 5 | 3c | 0.586 | derived | mortality | external model | |
| Congo, Democratic Republic of | COD | 5 | 3c | 0.481 | derived | mortality | external model | |
| Congo, Republic of | COG | 2b | 3c | 0.593 | derived | mortality | external model | |
| Costa Rica | CRI | 1 | 1 | 0.806 | independent | – | – | |
| Côte d'Ivoire | CIV | 4 | 3c | 0.534 | derived | mortality | external model | |
| Croatia | HRV | 1 | 1 | 0.878 | independent | – | – | |
| Cuba | CUB | 3c | 1 | 0.764 | derived | incidence | external model | |
| Curaçao | CUW | 2a | 3c | 0.946* | derived | mortality | external model | |
| Cyprus | CYP | 1 | 1 | 0.907 | independent | – | – | |
| Czechia | CZE | 1 | 1 | 0.895 | independent | – | – | |
| Denmark | DNK | 1 | 1 | 0.952 | independent | – | – | |
| Djibouti | DJI | 5 | 3c | 0.515 | derived | mortality | external model | |
| Dominican Republic | DOM | 3c | 4 | 0.766 | derived | incidence | external model | |
| Ecuador | ECU | 3c | 1 | 0.765 | derived | incidence | external model | |
| Egypt | EGY | 2b | 3c | 0.728 | derived | mortality | external model | |
| El Salvador | SLV | 3c | 1 | 0.674 | derived | incidence | external model | |
| Equatorial Guinea | GNQ | 5 | 3c | 0.650 | derived | mortality | external model | |
| Eritrea | ERI | 5 | 3c | 0.493 | derived | mortality | external model | |
| Estonia | EST | 1 | 1 | 0.899 | independent | – | – | |
| Eswatini | SWZ | 2a | 3c | 0.610 | derived | mortality | external model | |
| Ethiopia | ETH | 2b | 3c | 0.492 | derived | mortality | external model | |
| Fiji | FJI | 3b | 1 | 0.729 | derived | incidence | external model | Yes |
| Finland | FIN | 1 | 1 | 0.942 | independent | – | – | |
| France (metropolitan) | FRA | 3a | 1 | 0.910 | derived | incidence | own-data ratio | |

| Country | ISO3 | Incidence method | Mortality method | HDI | MIR class | Derived component | Ratio source | Method 3b |
|---|---|---|---|---|---|---|---|---|
| France, Guadeloupe | GLP | 1 | 3c | 0.910* | derived | mortality | external model | |
| France, La Réunion | REU | 2a | 1 | 0.910* | independent | – | – | |
| France, Martinique | MTQ | 1 | 1 | 0.910* | independent | – | – | |
| French Guyana | GUF | 3c | 1 | 0.910* | derived | incidence | external model | |
| French Polynesia | PYF | 1 | 3b | 0.910* | derived | mortality | external model | Yes |
| Gabon | GAB | 2b | 3c | 0.693 | derived | mortality | external model | |
| Gaza Strip and West Bank | PSE | 5 | 3c | 0.716 | derived | mortality | external model | |
| Georgia | GEO | 3c | 1 | 0.814 | derived | incidence | external model | |
| Germany | DEU | 1 | 1 | 0.950 | independent | – | – | |
| Ghana | GHA | 4 | 3c | 0.602 | derived | mortality | external model | |
| Greece | GRC | 3b | 1 | 0.893 | derived | incidence | external model | Yes |
| Guam | GUM | 5 | 3b | 0.927* | derived | mortality | external model | Yes |
| Guatemala | GTM | 3c | 1 | 0.629 | derived | incidence | external model | |
| Guinea | GIN | 2b | 3c | 0.471 | derived | mortality | external model | |
| Guinea-Bissau | GNB | 5 | 3c | 0.483 | derived | mortality | external model | |
| Guyana | GUY | 3c | 1 | 0.742 | derived | incidence | external model | |
| Haiti | HTI | 3c | 4 | 0.552 | derived | incidence | external model | |
| Honduras | HND | 2b | 3c | 0.624 | derived | mortality | external model | |
| Hungary | HUN | 3b | 1 | 0.851 | derived | incidence | external model | Yes |
| Iceland | ISL | 1 | 1 | 0.959 | independent | – | – | |
| India | IND | 2b | 3a | 0.644 | derived | mortality | own-data ratio | |
| Indonesia | IDN | 4 | 3c | 0.713 | derived | mortality | external model | |
| Iran, Islamic Republic of | IRN | 2a | 3c | 0.780 | derived | mortality | external model | |
| Iraq | IRQ | 2a | 3c | 0.673 | derived | mortality | external model | |
| Ireland | IRL | 1 | 1 | 0.950 | independent | – | – | |
| Israel | ISR | 1 | 1 | 0.915 | independent | – | – | |
| Italy | ITA | 3a | 1 | 0.906 | derived | incidence | own-data ratio | |
| Jamaica | JAM | 3c | 1 | 0.706 | derived | incidence | external model | |
| Japan | JPN | 1 | 1 | 0.920 | independent | – | – | |

| Country | ISO3 | Incidence method | Mortality method | HDI | MIR class | Derived component | Ratio source | Method 3b |
|---|---|---|---|---|---|---|---|---|
| Jordan | JOR | 3c | 1 | 0.736 | derived | incidence | external model | |
| Kazakhstan | KAZ | 3c | 1 | 0.802 | derived | incidence | external model | |
| Kenya | KEN | 2b | 3c | 0.601 | derived | mortality | external model | |
| Korea, Democratic People's Republic of | PRK | 5 | 3c | – | derived | mortality | external model | |
| Korea, Republic of | KOR | 1 | 1 | 0.929 | independent | – | – | |
| Kuwait | KWT | 2a | 3c | 0.847 | derived | mortality | external model | |
| Kyrgyzstan | KGZ | 3c | 1 | 0.701 | derived | incidence | external model | |
| Lao People's Democratic Republic | LAO | 5 | 3c | 0.620 | derived | mortality | external model | |
| Latvia | LVA | 1 | 1 | 0.879 | independent | – | – | |
| Lebanon | LBN | 2a | 3c | 0.723 | derived | mortality | external model | |
| Lesotho | LSO | 5 | 3c | 0.521 | derived | mortality | external model | |
| Liberia | LBR | 5 | 3c | 0.487 | derived | mortality | external model | |
| Libya | LBY | 2b | 3c | 0.746 | derived | mortality | external model | |
| Lithuania | LTU | 1 | 1 | 0.879 | independent | – | – | |
| Luxembourg | LUX | 3b | 1 | 0.927 | derived | incidence | external model | Yes |
| Madagascar | MDG | 5 | 3c | 0.487 | derived | mortality | external model | |
| Malawi | MWI | 2b | 3c | 0.508 | derived | mortality | external model | |
| Malaysia | MYS | 2b | 3c | 0.807 | derived | mortality | external model | |
| Maldives | MDV | 5 | 3c | 0.762 | derived | mortality | external model | |
| Mali | MLI | 2b | 3c | 0.410 | derived | mortality | external model | |
| Malta | MLT | 1 | 1 | 0.915 | independent | – | – | |
| Mauritania | MRT | 5 | 3c | 0.540 | derived | mortality | external model | |
| Mauritius | MUS | 2a | 1 | 0.796 | independent | – | – | |
| Mexico | MEX | 3b | 1 | 0.781 | derived | incidence | external model | Yes |
| Mongolia | MNG | 3c | 1 | 0.741 | derived | incidence | external model | |
| Montenegro | MNE | 3b | 2a | 0.844 | derived | incidence | external model | Yes |
| Morocco | MAR | 2b | 3c | 0.698 | derived | mortality | external model | |
| Mozambique | MOZ | 2b | 3c | 0.461 | derived | mortality | external model | |
| Myanmar | MMR | 2b | 3c | 0.608 | derived | mortality | external model | |

| Country | ISO3 | Incidence method | Mortality method | HDI | MIR class | Derived component | Ratio source | Method 3b |
|---|---|---|---|---|---|---|---|---|
| Namibia | NAM | 2a | 3c | 0.610 | derived | mortality | external model | |
| Nepal | NPL | 5 | 3c | 0.601 | derived | mortality | external model | |
| New Caledonia | NCL | 1 | 3b | 0.910* | derived | mortality | external model | Yes |
| New Zealand | NZL | 1 | 1 | 0.939 | independent | – | – | |
| Nicaragua | NIC | 3c | 1 | 0.669 | derived | incidence | external model | |
| Niger | NER | 4 | 3c | 0.394 | derived | mortality | external model | |
| Nigeria | NGA | 4 | 3c | 0.548 | derived | mortality | external model | |
| North Macedonia | MKD | 3b | 1 | 0.765 | derived | incidence | external model | Yes |
| Norway | NOR | 1 | 1 | 0.966 | independent | – | – | |
| Oman | OMN | 2a | 3c | 0.819 | derived | mortality | external model | |
| Pakistan | PAK | 4 | 3c | 0.540 | derived | mortality | external model | |
| Panama | PAN | 3c | 1 | 0.820 | derived | incidence | external model | |
| Papua New Guinea | PNG | 3c | 4 | 0.568 | derived | incidence | external model | |
| Paraguay | PRY | 3c | 1 | 0.731 | derived | incidence | external model | |
| Peru | PER | 3a | 1 | 0.762 | derived | incidence | own-data ratio | |
| Philippines | PHL | 2b | 1 | 0.710 | independent | – | – | |
| Poland | POL | 1 | 1 | 0.881 | independent | – | – | |
| Portugal | PRT | 3a | 1 | 0.874 | derived | incidence | own-data ratio | |
| Puerto Rico | PRI | 1 | 3c | 0.927* | derived | mortality | external model | |
| Qatar | QAT | 2a | 3c | 0.875 | derived | mortality | external model | |
| Republic of Moldova | MDA | 3b | 1 | 0.763 | derived | incidence | external model | Yes |
| Romania | ROU | 3b | 1 | 0.827 | derived | incidence | external model | Yes |
| Russian Federation | RUS | 2b | 1 | 0.821 | independent | – | – | |
| Rwanda | RWA | 2b | 3c | 0.548 | derived | mortality | external model | |
| Saint Lucia | LCA | 3c | 1 | 0.725 | derived | incidence | external model | |
| Samoa | WSM | 5 | 3b | 0.702 | derived | mortality | external model | Yes |
| Sao Tome and Principe | STP | 5 | 3c | 0.613 | derived | mortality | external model | |
| Saudi Arabia | SAU | 2a | 3c | 0.875 | derived | mortality | external model | |
| Senegal | SEN | 5 | 3c | 0.517 | derived | mortality | external model | |

| Country | ISO3 | Incidence method | Mortality method | HDI | MIR class | Derived component | Ratio source | Method 3b |
|---|---|---|---|---|---|---|---|---|
| Serbia | SRB | 3b | 1 | 0.805 | derived | incidence | external model | Yes |
| Sierra Leone | SLE | 5 | 3c | 0.458 | derived | mortality | external model | |
| Singapore | SGP | 3a | 1 | 0.949 | derived | incidence | own-data ratio | |
| Slovakia | SVK | 3a | 1 | 0.855 | derived | incidence | own-data ratio | |
| Slovenia | SVN | 1 | 1 | 0.926 | independent | – | – | |
| Solomon Islands | SLB | 5 | 3c | 0.562 | derived | mortality | external model | |
| Somalia | SOM | 5 | 3c | 0.380 | derived | mortality | external model | |
| South Africa | ZAF | 4 | 1 | 0.717 | separate | – | – | |
| South Sudan | SSD | 5 | 3c | 0.381 | derived | mortality | external model | |
| Spain | ESP | 3a | 1 | 0.911 | derived | incidence | own-data ratio | |
| Sri Lanka | LKA | 2b | 3c | 0.780 | derived | mortality | external model | |
| Sudan | SDN | 4 | 3c | 0.516 | derived | mortality | external model | |
| Suriname | SUR | 3c | 1 | 0.690 | derived | incidence | external model | |
| Sweden | SWE | 1 | 1 | 0.952 | independent | – | – | |
| Switzerland | CHE | 3a | 1 | 0.967 | derived | incidence | own-data ratio | |
| Syrian Arab Republic | SYR | 5 | 3c | 0.557 | derived | mortality | external model | |
| Tajikistan | TJK | 3c | 2a | 0.679 | derived | incidence | external model | |
| Tanzania, United Republic of | TZA | 2b | 3c | 0.532 | derived | mortality | external model | |
| Thailand | THA | 2b | 3c | 0.803 | derived | mortality | external model | |
| The Netherlands | NLD | 1 | 1 | 0.946 | independent | – | – | |
| The Republic of the Gambia | GMB | 4 | 3c | 0.495 | derived | mortality | external model | |
| Timor-Leste | TLS | 5 | 3c | 0.566 | derived | mortality | external model | |
| Togo | TGO | 5 | 3c | 0.547 | derived | mortality | external model | |
| Trinidad and Tobago | TTO | 2a | 1 | 0.814 | independent | – | – | |
| Tunisia | TUN | 2b | 3c | 0.732 | derived | mortality | external model | |
| Türkiye | TUR | 2b | 3a | 0.855 | derived | mortality | own-data ratio | |
| Turkmenistan | TKM | 3c | 1 | 0.744 | derived | incidence | external model | |
| Uganda | UGA | 2b | 3c | 0.550 | derived | mortality | external model | |
| Ukraine | UKR | 1 | 1 | 0.734 | independent | – | – | |

| Country | ISO3 | Incidence method | Mortality method | HDI | MIR class | Derived component | Ratio source | Method 3b |
|---|---|---|---|---|---|---|---|---|
| United Arab Emirates | ARE | 2a | 3c | 0.937 | derived | mortality | external model | |
| United Kingdom | GBR | 1 | 1 | 0.940 | independent | – | – | |
| United States of America | USA | 1 | 1 | 0.927 | independent | – | – | |
| Uruguay | URY | 1 | 1 | 0.830 | independent | – | – | |
| Uzbekistan | UZB | 3c | 1 | 0.727 | derived | incidence | external model | |
| Vanuatu | VUT | 5 | 3b | 0.614 | derived | mortality | external model | Yes |
| Venezuela | VEN | 3b | 1 | 0.699 | derived | incidence | external model | Yes |
| Viet Nam | VNM | 2b | 3c | 0.726 | derived | mortality | external model | |
| Yemen | YEM | 5 | 3c | 0.424 | derived | mortality | external model | |
| Zambia | ZMB | 2b | 3c | 0.569 | derived | mortality | external model | |
| Zimbabwe | ZWE | 2b | 3c | 0.550 | derived | mortality | external model | |

*NOTE. Independent MIR: both components observed (methods 1, 2a, or 2b) and neither derived from the other. Derived MIR: at least one component derived from the other; "derived component" names the component that was derived. Ratio source: own-data ratios (method 3a) or external models (methods 3b, 3c, or regional/neighboring estimates). Method 3b: HDI-scaled ratios for incidence or mortality. HDI values are from the Human Development Report 2023-2024; *territories assigned the HDI of their administering country.*

***Supplementary Table S2.*** *Change in Spearman Correlation Between MIR and HDI Under Four Restriction Schemes*

| Cancer site | n | ρ (full) | A n | A Δρ (95% CI) | B n | B Δρ (95% CI) | C n | C Δρ (95% CI) | D n | D Δρ (95% CI) |
|---|---|---|---|---|---|---|---|---|---|---|
| Cervix uteri | 151 | −0.88 | 73 | 0.001 (−0.03 to 0.05) | 76 | 0.077 (0.01 to 0.17) | 35 | 0.165 (0.04 to 0.36) | 141 | 0.003 (−0.01 to 0.02) |
| Testis | 84 | −0.88 | 47 | −0.024 (−0.08 to 0.04) | 57 | 0.016 (−0.05 to 0.09) | 30 | 0.004 (−0.10 to 0.16) | 78 | −0.002 (−0.02 to 0.01) |
| Non-Hodgkin lymphoma | 144 | −0.86 | 75 | −0.003 (−0.05 to 0.05) | 73 | 0.039 (−0.02 to 0.12) | 36 | 0.049 (−0.07 to 0.25) | 135 | −0.002 (−0.01 to 0.01) |
| Prostate | 171 | −0.85 | 86 | −0.025 (−0.08 to 0.04) | 88 | 0.076 (0.02 to 0.15) | 41 | 0.138 (−0.02 to 0.37) | 157 | −0.001 (−0.01 to 0.01) |
| Leukemia | 141 | −0.85 | 72 | −0.022 (−0.07 to 0.04) | 73 | 0.000 (−0.07 to 0.08) | 34 | 0.108 (−0.05 to 0.35) | 132 | −0.001 (−0.01 to 0.01) |
| Breast | 173 | −0.84 | 87 | 0.009 (−0.05 to 0.09) | 86 | 0.019 (−0.06 to 0.10) | 41 | 0.077 (−0.05 to 0.27) | 159 | 0.004 (−0.01 to 0.02) |
| Lip, oral cavity | 119 | −0.83 | 65 | 0.013 (−0.04 to 0.08) | 63 | 0.084 (0.00 to 0.19) | 33 | 0.078 (−0.04 to 0.26) | 111 | −0.006 (−0.02 to 0.01) |
| Bladder | 136 | −0.83 | 72 | 0.036 (−0.01 to 0.11) | 73 | 0.075 (−0.02 to 0.19) | 36 | 0.187 (0.01 to 0.42) | 125 | −0.001 (−0.02 to 0.02) |
| Hodgkin lymphoma | 98 | −0.83 | 54 | −0.017 (−0.08 to 0.06) | 54 | −0.038 (−0.12 to 0.04) | 26 | 0.068 (−0.10 to 0.35) | 92 | −0.006 (−0.03 to 0.01) |
| Salivary glands | 66 | −0.83 | 37 | −0.002 (−0.09 to 0.11) | 38 | 0.273 (0.13 to 0.48) | 19 | 0.246 (−0.00 to 0.66) | 60 | −0.018 (−0.05 to 0.00) |
| Thyroid | 130 | −0.82 | 66 | 0.124 (0.03 to 0.26) | 72 | 0.222 (0.11 to 0.37) | 33 | 0.818 (0.47 to 1.19) | 121 | −0.013 (−0.03 to 0.00) |
| Lung | 154 | −0.82 | 80 | 0.042 (−0.02 to 0.13) | 82 | 0.031 (−0.04 to 0.11) | 40 | 0.234 (0.05 to 0.46) | 141 | −0.004 (−0.03 to 0.02) |
| Corpus uteri | 133 | −0.81 | 68 | 0.023 (−0.05 to 0.12) | 75 | 0.113 (0.03 to 0.22) | 36 | 0.357 (0.12 to 0.67) | 123 | −0.015 (−0.04 to 0.00) |
| Stomach | 144 | −0.80 | 74 | −0.016 (−0.08 to 0.06) | 75 | −0.051 (−0.13 to 0.03) | 36 | 0.054 (−0.08 to 0.25) | 135 | 0.004 (−0.01 to 0.02) |
| Colon | 154 | −0.80 | 81 | 0.012 (−0.04 to 0.09) | 83 | −0.008 (−0.07 to 0.06) | 41 | 0.023 (−0.07 to 0.16) | 143 | 0.005 (−0.01 to 0.02) |
| Melanoma of skin | 96 | −0.79 | 53 | −0.016 (−0.09 to 0.08) | 64 | 0.050 (−0.03 to 0.16) | 34 | −0.005 (−0.12 to 0.14) | 85 | −0.014 (−0.04 to 0.01) |
| Brain, CNS | 129 | −0.78 | 67 | −0.040 (−0.13 to 0.07) | 71 | 0.069 (−0.05 to 0.18) | 34 | 0.322 (0.08 to 0.62) | 120 | −0.004 (−0.03 to 0.01) |
| Larynx | 107 | −0.76 | 58 | −0.049 (−0.12 to 0.03) | 61 | 0.040 (−0.05 to 0.14) | 31 | 0.073 (−0.10 to 0.37) | 98 | 0.002 (−0.01 to 0.02) |
| Kidney | 129 | −0.75 | 67 | 0.048 (−0.02 to 0.14) | 72 | 0.007 (−0.09 to 0.12) | 34 | 0.298 (0.07 to 0.60) | 120 | 0.007 (−0.00 to 0.02) |
| Vulva | 65 | −0.75 | 38 | 0.050 (−0.04 to 0.19) | 41 | 0.018 (−0.11 to 0.15) | 23 | 0.011 (−0.18 to 0.32) | 59 | 0.012 (−0.03 to 0.06) |
| Multiple myeloma | 98 | −0.75 | 57 | −0.040 (−0.12 to 0.04) | 58 | −0.042 (−0.14 to 0.06) | 33 | −0.025 (−0.18 to 0.20) | 92 | 0.008 (−0.01 to 0.03) |
| Nasopharynx | 68 | −0.74 | 32 | 0.081 (−0.08 to 0.33) | 29 | 0.245 (0.02 to 0.54) | 13 | 0.264 (−0.06 to 0.87) | 64 | −0.007 (−0.05 to 0.03) |
| Other specified cancers | 157 | −0.73 | 81 | −0.014 (−0.07 to 0.06) | 80 | 0.043 (−0.08 to 0.18) | 40 | 0.067 (−0.11 to 0.27) | 147 | 0.007 (−0.01 to 0.03) |
| Anus | 65 | −0.73 | 39 | −0.038 (−0.16 to 0.10) | 38 | 0.274 (0.08 to 0.51) | 21 | 0.036 (−0.18 to 0.36) | 60 | −0.039 (−0.11 to 0.01) |
| Ovary | 136 | −0.72 | 69 | 0.093 (−0.01 to 0.22) | 73 | 0.274 (0.11 to 0.46) | 35 | 0.707 (0.34 to 1.07) | 127 | −0.012 (−0.03 to 0.00) |
| Penis | 44 | −0.72 | 26 | −0.046 (−0.19 to 0.13) | 29 | 0.158 (−0.01 to 0.39) | 16 | −0.043 (−0.26 to 0.30) | 40 | 0.009 (−0.03 to 0.05) |

| Cancer site | n | ρ (full) | A n | A Δρ (95% CI) | B n | B Δρ (95% CI) | C n | C Δρ (95% CI) | D n | D Δρ (95% CI) |
|---|---|---|---|---|---|---|---|---|---|---|
| Oropharynx | 69 | −0.71 | 38 | −0.046 (−0.16 to 0.09) | 48 | 0.111 (0.00 to 0.26) | 27 | 0.024 (−0.16 to 0.30) | 63 | −0.034 (−0.11 to 0.01) |
| Gallbladder | 73 | −0.71 | 40 | 0.007 (−0.13 to 0.16) | 41 | 0.215 (0.02 to 0.43) | 22 | 0.247 (−0.01 to 0.59) | 68 | 0.009 (−0.01 to 0.03) |
| Kaposi sarcoma | 43 | −0.70 | 21 | 0.069 (−0.11 to 0.38) | 19 | 0.324 (−0.08 to 0.84) | 7 | 0.661 (−0.38 to 1.60) | 40 | 0.032 (−0.00 to 0.11) |
| Rectum | 141 | −0.69 | 74 | 0.019 (−0.08 to 0.13) | 74 | 0.173 (0.06 to 0.31) | 36 | 0.167 (−0.04 to 0.43) | 131 | −0.008 (−0.03 to 0.01) |
| Pancreas | 138 | −0.65 | 72 | 0.015 (−0.08 to 0.12) | 77 | −0.036 (−0.19 to 0.13) | 38 | 0.277 (−0.02 to 0.58) | 128 | 0.019 (0.00 to 0.05) |
| Esophagus | 115 | −0.64 | 59 | −0.007 (−0.11 to 0.11) | 61 | −0.019 (−0.19 to 0.16) | 31 | 0.132 (−0.14 to 0.44) | 109 | 0.005 (−0.02 to 0.03) |
| Vagina | 32 | −0.57 | 18 | −0.092 (−0.33 to 0.22) | 21 | 0.673 (0.29 to 1.08) | 12 | 0.421 (−0.19 to 1.10) | 30 | −0.046 (−0.14 to 0.00) |
| Unspecified sites | 140 | −0.54 | 70 | 0.255 (0.11 to 0.42) | 75 | 0.549 (0.38 to 0.73) | 36 | 0.841 (0.55 to 1.08) | 130 | −0.008 (−0.03 to 0.01) |
| Liver | 148 | −0.52 | 72 | 0.005 (−0.10 to 0.13) | 73 | −0.174 (−0.33 to 0.01) | 33 | 0.004 (−0.30 to 0.37) | 138 | −0.016 (−0.06 to 0.02) |
| Hypopharynx | 56 | −0.50 | 32 | 0.011 (−0.17 to 0.22) | 37 | 0.171 (−0.01 to 0.40) | 21 | −0.049 (−0.33 to 0.35) | 52 | −0.013 (−0.05 to 0.01) |
| Mesothelioma | 34 | −0.04 | 23 | 0.105 (−0.24 to 0.47) | 28 | 0.269 (0.03 to 0.57) | 19 | 0.260 (−0.20 to 0.72) | 33 | −0.021 (−0.08 to 0.00) |

*NOTE. Δρ = ρ_restricted − ρ_full; for negative correlations, a positive Δρ means a weaker (less negative) correlation. 95% CIs are from a paired bootstrap (2,000 resamples of countries). Scheme A, registry-based incidence (methods 1, 2a, 2b); Scheme B, observed mortality (methods 1, 2a, 2b); Scheme C, independent MIR (both components observed, neither derived from the other); Scheme D, exclusion of the 17 countries using method 3b for incidence or mortality. n, number of countries with at least 100 estimated cases and nonzero age-standardized incidence. Sites are ordered by ascending full-sample ρ.*

***Supplementary Table S3.*** *HDI-Matched Subsampling Results for the Independent-MIR Restriction (Scheme C)*

| Cancer site | n (full) | ρ (full) | n (C) | ρ (C) | Null median | Null 95% interval | One-sided p | BH-adjusted p |
|---|---|---|---|---|---|---|---|---|
| Testis | 84 | −0.88 | 30 | −0.874 | −0.88 | −0.93 to −0.80 | 0.492 | 0.908 |
| Non-Hodgkin lymphoma | 144 | −0.86 | 36 | −0.814 | −0.73 | −0.84 to −0.61 | 0.927 | 0.953 |
| Melanoma of skin | 96 | −0.79 | 34 | −0.795 | −0.73 | −0.84 to −0.59 | 0.862 | 0.938 |
| Colon | 154 | −0.80 | 41 | −0.773 | −0.73 | −0.81 to −0.62 | 0.837 | 0.938 |
| Multiple myeloma | 98 | −0.75 | 33 | −0.772 | −0.73 | −0.83 to −0.63 | 0.768 | 0.938 |
| Hodgkin lymphoma | 98 | −0.83 | 26 | −0.763 | −0.81 | −0.91 to −0.71 | 0.222 | 0.588 |
| Breast | 173 | −0.84 | 41 | −0.760 | −0.68 | −0.82 to −0.55 | 0.842 | 0.938 |
| Penis | 44 | −0.72 | 16 | −0.758 | −0.66 | −0.81 to −0.36 | 0.892 | 0.943 |
| Lip, oral cavity | 119 | −0.83 | 33 | −0.755 | −0.66 | −0.76 to −0.53 | 0.963 | 0.963 |
| Vulva | 65 | −0.75 | 23 | −0.743 | −0.79 | −0.88 to −0.66 | 0.209 | 0.588 |
| Stomach | 144 | −0.80 | 36 | −0.742 | −0.73 | −0.83 to −0.62 | 0.589 | 0.908 |
| Leukemia | 141 | −0.85 | 34 | −0.738 | −0.73 | −0.84 to −0.60 | 0.572 | 0.908 |
| Prostate | 171 | −0.85 | 41 | −0.715 | −0.72 | −0.81 to −0.62 | 0.449 | 0.874 |
| Cervix uteri | 151 | −0.88 | 35 | −0.714 | −0.77 | −0.86 to −0.65 | 0.171 | 0.575 |
| Anus | 65 | −0.73 | 21 | −0.697 | −0.59 | −0.80 to −0.32 | 0.790 | 0.938 |
| Larynx | 107 | −0.76 | 31 | −0.687 | −0.74 | −0.82 to −0.65 | 0.110 | 0.428 |
| Oropharynx | 69 | −0.71 | 27 | −0.685 | −0.63 | −0.75 to −0.44 | 0.785 | 0.938 |
| Other specified cancers | 157 | −0.73 | 40 | −0.667 | −0.62 | −0.75 to −0.46 | 0.743 | 0.938 |
| Bladder | 136 | −0.83 | 36 | −0.644 | −0.64 | −0.77 to −0.48 | 0.527 | 0.908 |
| Salivary glands | 66 | −0.83 | 19 | −0.583 | −0.64 | −0.83 to −0.37 | 0.346 | 0.758 |
| Lung | 154 | −0.82 | 40 | −0.582 | −0.70 | −0.81 to −0.56 | 0.053 | 0.246 |
| Hypopharynx | 56 | −0.50 | 21 | −0.552 | −0.42 | −0.65 to −0.16 | 0.848 | 0.938 |
| Rectum | 141 | −0.69 | 36 | −0.525 | −0.47 | −0.65 to −0.30 | 0.735 | 0.938 |
| Liver | 148 | −0.52 | 33 | −0.518 | −0.58 | −0.74 to −0.39 | 0.240 | 0.593 |
| Esophagus | 115 | −0.64 | 31 | −0.506 | −0.57 | −0.74 to −0.40 | 0.215 | 0.588 |
| Nasopharynx | 68 | −0.74 | 13 | −0.478 | −0.53 | −0.77 to −0.26 | 0.353 | 0.758 |
| Gallbladder | 73 | −0.71 | 22 | −0.458 | −0.60 | −0.77 to −0.36 | 0.116 | 0.428 |
| Brain, CNS | 129 | −0.78 | 34 | −0.456 | −0.63 | −0.77 to −0.47 | 0.019 | 0.222 |
| Kidney | 129 | −0.75 | 34 | −0.456 | −0.61 | −0.73 to −0.46 | 0.024 | 0.222 |
| Corpus uteri | 133 | −0.81 | 36 | −0.449 | −0.58 | −0.71 to −0.42 | 0.050 | 0.246 |
| Pancreas | 138 | −0.65 | 38 | −0.373 | −0.54 | −0.70 to −0.36 | 0.037 | 0.226 |
| Vagina | 32 | −0.57 | 12 | −0.147 | −0.09 | −0.47 to 0.32 | 0.620 | 0.918 |
| Kaposi sarcoma | 43 | −0.70 | 7 | −0.036 | −0.14 | −0.79 to 0.46 | 0.369 | 0.758 |
| Ovary | 136 | −0.72 | 35 | −0.016 | −0.35 | −0.56 to −0.12 | 0.003 | 0.056 |
| Thyroid | 130 | −0.82 | 33 | −0.004 | −0.35 | −0.53 to −0.15 | <0.001 | **0.011** |
| Mesothelioma | 34 | −0.04 | 19 | 0.216 | 0.23 | −0.08 to 0.54 | 0.543 | 0.908 |
| Unspecified sites | 140 | −0.54 | 36 | 0.300 | 0.10 | −0.13 to 0.31 | 0.031 | 0.226 |

*NOTE. For each site, 10,000 random subsets of the full sample were drawn without replacement, matching the Scheme C distribution across HDI quartiles. One-sided p = (k + 1)/(N + 1), where k is the number of subsets with a correlation at least as attenuated as observed. BH, Benjamini-Hochberg adjustment across 37 sites; bold, BH-adjusted p < 0.05. Sites are ordered by Scheme C ρ.*
